# Hot days and Covid-19 – unusual heat stress for nursing professions in Germany

**DOI:** 10.1101/2021.01.29.21250592

**Authors:** Yvette Jegodka, Lena Lagally, Hanna Mertes, Katharina Deering, Julia Schoierer, Barbara Buchberger, Stephan Bose-O’Reilly

## Abstract

**Objectives:** Our aim was to identify whether working during hot days alongside with Covid-19 related personal protective equipment causes heat stress for nursing professionals in Germany.

**Methods:** Using an online survey, we assessed the impact of hot weather on nursing staff performing in personal protective equipment. A random selection of nursing staff from hospitals, nursing homes and outpatient care participated in the survey.

**Results:** Out of 428 participants, 6.3% were between 16 and 25 years old, 22.8% between 26 and 35 years, 21.9% between 36 and 45 years, 30.5% between 45 and 55 years, 18.2% between 56 and 65 years, and 0.3% were older than 65 years. Out of all participants, 18.2% were male and 82.5% female. The results of the survey showed that 48.3% had more than 20 years of experience in nursing and 46.2% cardiac, pulmonary, or other pre-existing conditions. Work was found exhaustive while working in PPE by 96.5% of the participants, and 93% complained of worse breathing. We found out that 85.8% reported difficulties to focus. Many workplaces turned out to lack adequate heat protection, with distinct differences concerning the amount of prophylactic and heat mitigating measures across institutions.

**Conclusions:** Our results clearly show that employers must make more of an effort to provide adequate heat protection for their nursing staff. In order to secure the public health care, there is a need for action, especially in the case of previous conditions of caregivers.

**What is already known about this subject?:**

► Working in personal protective equipment is often needed during pandemics, to protect nurses, doctors and staff from an infection.
► However, the equipment can also hamper efficiency and productivity of healthcare workers and lead to personal discomfort, for example, during heat waves.

**What are the new findings?:**

► According to our study, nurses and nursing assistants in Germany are often older than 45 years and, in many cases, suffer from pre-existing conditions, which exacerbate the problems with personal protective equipment during periods of hot temperatures.
► Many healthcare institutions do not offer adequate ways to mitigate heat stress for their staff.

**How might this impact on policy or clinical practice in the foreseeable future?:**

► The results from this study can inform policy makers and clinical practitioners to modify their protocols to include better protective measures during extreme heat or other adverse environmental conditions.

## Introduction

One of the greatest health threats in our century is climate change [1]. The Intergovernmental Panel on Climate Change reports a clear increase in global warming, which negatively affects human health [2]. This increase will both concern average temperatures as well as the number of heat wave spikes [3]. A recent study compared heat stress experiences of nurses in personal protective equipment (PPE) working in India and Singapore during the 2020 Covid-19 pandemic [4,5]. Healthcare workers from both countries reported a high degree of thirst, sweating, exhaustion and an increased desire to move into comfort zones. Singaporean nurses had more choices to mitigate thermal stress in form of air conditioning, available rest areas and the opportunity to take off PPE during breaks [5] as the study found. Healthcare workers are often aware of their situation, but with few opportunities for relief due to work-based constraints [6]. If institutional environments do not provide dedicated measures to mitigate heat stress, nurses may not take evasive measures themselves [5].

According to the World Health Organization (WHO), one lesson learned from the West-African Ebola disease outbreaks of the last decade was that “personal protective equipment is hot and cumbersome” [7,8]. However, in the case of Ebola, heat stress can be mitigated by including ventilation within the PPE, as Ebola is not an airborne disease [8]. This option, of course, is not available for handling SARS-CoV-2, as the latter can indeed be transmitted through droplets [9]. Daanen et al. (2020) suggest the implementation of strategic measures - adjusting work and rest times, wearing lighter clothing, and drinking cold water to precool and to reduce the increase in body core temperature [10].

In August 2020, many parts across Germany experienced a three-week long heat wave. The double burden of infection protection measures due to the SARS-CoV-2 pandemic situation and exposure to heat may have led to a particular challenge for nurses, as it is known that wearing PPE alone can lead to headaches and other mental and physical symptoms [11]. Existing heat-health plans may include a clearly outlined response to heat emergencies, a timely alert system, and a reduction in exposure to indoor heat [12], [13].

Importantly, according to the Federal Statistical Office of Germany, there were 6% excess deaths during August 2020 because of the extreme heat wave in the country. During the week of August 10^th^, the number was even 20% higher than the average of the same week between 2016 and 2019 [14]. This stresses once again that protecting workers in PPE from heat stress is an important goal even without the presence of a pandemic [15,16].

### Objectives

The aim was to identify whether working during hot days alongside with Corona virus disease (COVID-19) related personal protective equipment causes heat stress for nursing professions in Germany.

Central questions of this study were:

- Are caregivers exposed to increased heat stress by working in PPE on hot days?
- What measures are being taken to better protect them from heat strain?
- Are there differences in behavioural and condition-oriented prevention when comparing nursing staff in hospitals, nursing homes and outpatient care services?
- What are the implications of wearing personal protective equipment, due to the ongoing pandemic, and working on very hot days, for nursing professions?

## Methods

We collected data for this study via an online questionnaire. The construction of this survey and the discussion of the results were supported by a literature search using the Pubmed, Google Scholar and ResearchGate databases. Due to the novelty of the subject we also took, non-peer reviewed, ‘grey’ literature into account.

### Study procedure and design

The study was a standardized, descriptive cross-sectional survey. It collected structural data, such as participants’ years of employment, area of work; their usage of personal protective equipment (PPE); their experiences and perspectives in relation to the workload and job satisfaction on hot days; as well as their personal feelings of security. If there were more than two answering possibilities, a 4-point Likert scale was used. The options were ‘yes’, ‘likely’, ‘unlikely’, ‘no’. The questionnaire was created using LimeSurvey and was open to study participants online from the 1^st^ to the 31^st^ of August 2020. In total, the survey contained 66 questions that could be answered roughly within 20 minutes.

### Study participants

Participants were nurses and nursing assistants from various healthcare homes, hospitals and ambulatory care associations in Germany. Test subjects were asked to participate via personal communication, email and various social media channels. An information letter was used to further educate the participants about the nature of the study. In addition, participants were encouraged to notify colleagues about the study, allowing the number of people who took the study to snowball.

### Ethical considerations

The data collection was based exclusively on prospectively anonymised questionnaires. To guarantee privacy, no personally identifiable data was requested or recorded. Participation in the survey was voluntary. Participants had to give informed consent before engaging, by ticking a button within this prospectively anonymised questionnaire, that they agree to participate. Their anonymity, their right to cancel participation and the protection of people from harm were guaranteed. [17]. According to the standards of the ethical committee of the medical faculty at our university, LMU, no ethical clearance is needed for such a research, built on prospectively anonymised questionnaires. The Ethics Commission of the Medical Faculty LMU has released a corresponding waiver for this project (21-0103 KB).

### Statistical methods

Completely and incompletely answered questionnaires were analysed using both, Chi-square tests for independence and Fisher’s exact tests (SPSS version 24). Graphics and diagrams were created using R 4.0.

## Results

A total of 428 nurses took part in the survey, 254 nurses worked in hospitals, 101 in nursing homes and 73 in outpatient care (table S1). We found that 82.4% were women, 30.5% were between 45 and 55 years old, and 18.5% were older than 55 years. We identified 61.9% of participants working full-time, 37.2% part-time and 0.9% had a mini-job. Almost 70% of those questioned had been employed in the healthcare sector in Germany for many years. Of those questioned, 48.3% had worked in the profession for more than 20 years, and 20.9% had worked there for 11-20 years. We identified that 91.3% of all respondents found their job strenuous and 80.2% suffered from physical complaints such as back pain, sleep disorders, exhaustion or headaches. An amount of 46.2% had a previous illness suffering from hypertension, mental illness, diabetes, asthma, COPD or skin diseases. It was 59.2% of those questioned who felt that their work was not appreciated. In outpatient care 91.7% of the employees worked alone, 40.6% in a nursing home and 24.4% in a hospital. Not having not enough protective clothing in their workplace was reported by 30.2% of the nursing staff employed in the clinic. In the nursing homes and outpatient care services it was 32.7% and 32.9%.

### Consequences of working in personal protective equipment on hot days

When working in PPE, nearly all (99.5%) respondents said that they were sweating more, and 93.0% had trouble breathing. 88.6% needed more time to work, 85.8% found it harder to concentrate, and 49.9% said they were afraid of making mistakes (figure 1). 71.6% were on their own when it came to moving patients, even those with a higher bodyweight. For 57.6%, the necessity of frequently changing PPE was particularly stressful. This was the case when there was not enough nursing staff to assist with the work in the isolation room, and when the nursing staff had to procure missing material outside the isolation room.

**Figure 1.**
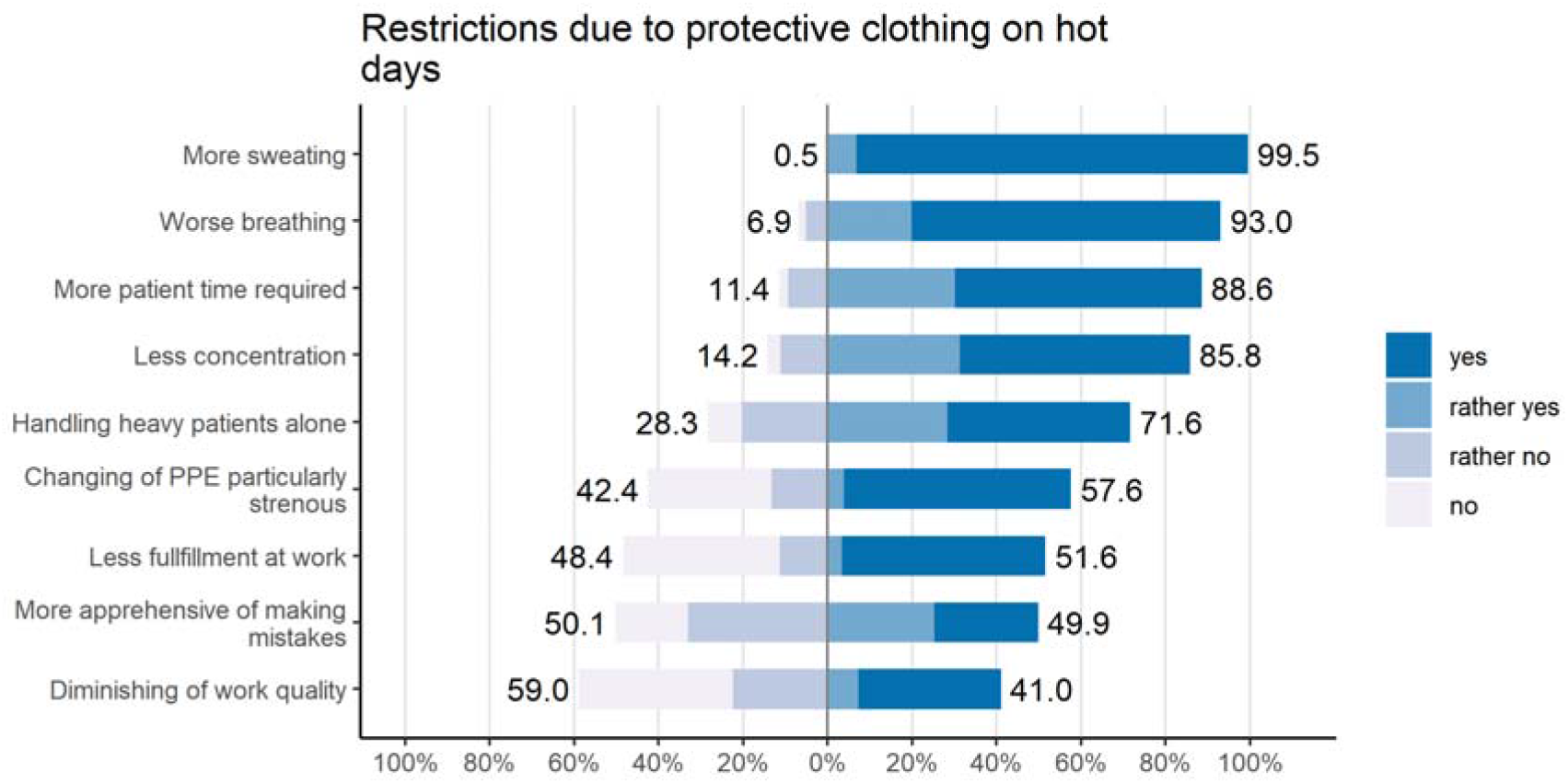
Restrictions due to protective clothing on hot days.

### Physical and psychological complaints when working in PPE during hot days

Heat exposure had an impact on emotional experience and physical well-being. A significant proportion of those questioned reported exhaustion and irritability. When working in protective clothing on hot days, 96.5% said they were particularly exhausted, 90.3% complained of tiredness, 82.8% were dissatisfied, 74.8% irritated. A majority of 71.2% of the participants suffered from headaches, and shortness of breath was a problem for 69.3%. Skin Problems were reported in 57.8% of individuals participating in the study (figure 2).

**Figure 2.**
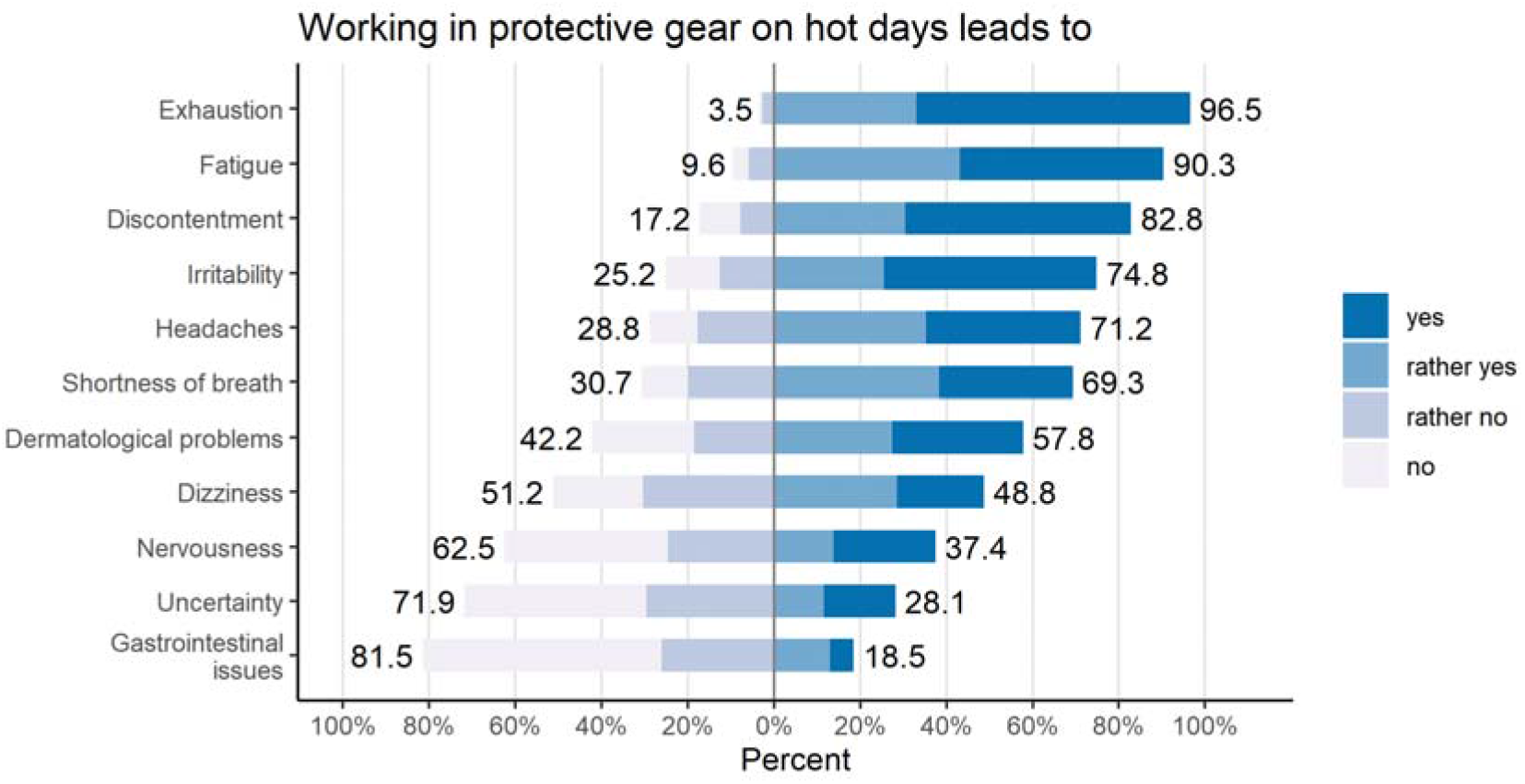
Consequences of working in PPE on hot days.

### Short-term countermeasures to prevent heat problems when working in PPE (behavioural prevention)

In order to avoid adverse effects from heat, the nursing staff took certain countermeasures when working in PPE on hot days. Most commonly reported was an increase in liquid uptake (67.0%), likely in order to compensate for the loss of fluid due to sweating. More drinks were only available for 42.9% of the survey participants, and 73.9% did not have enough colleagues to help them. A majority of 96.9% of individuals surveyed mentioned that no additional nursing staff was hired (figure 3). Only for 8.7% of employees it was possible to take more and/or longer breaks in order to better allocate their efforts or to regenerate more quickly. Just 9.2% were able to make changes to their care planning, and 9.1% could do strenuous work at cooler times of the day. Shorter workdays were only possible in 6.3% of the cases in outpatient care (figure 3, table S2, table S3,).

**Figure 3.**
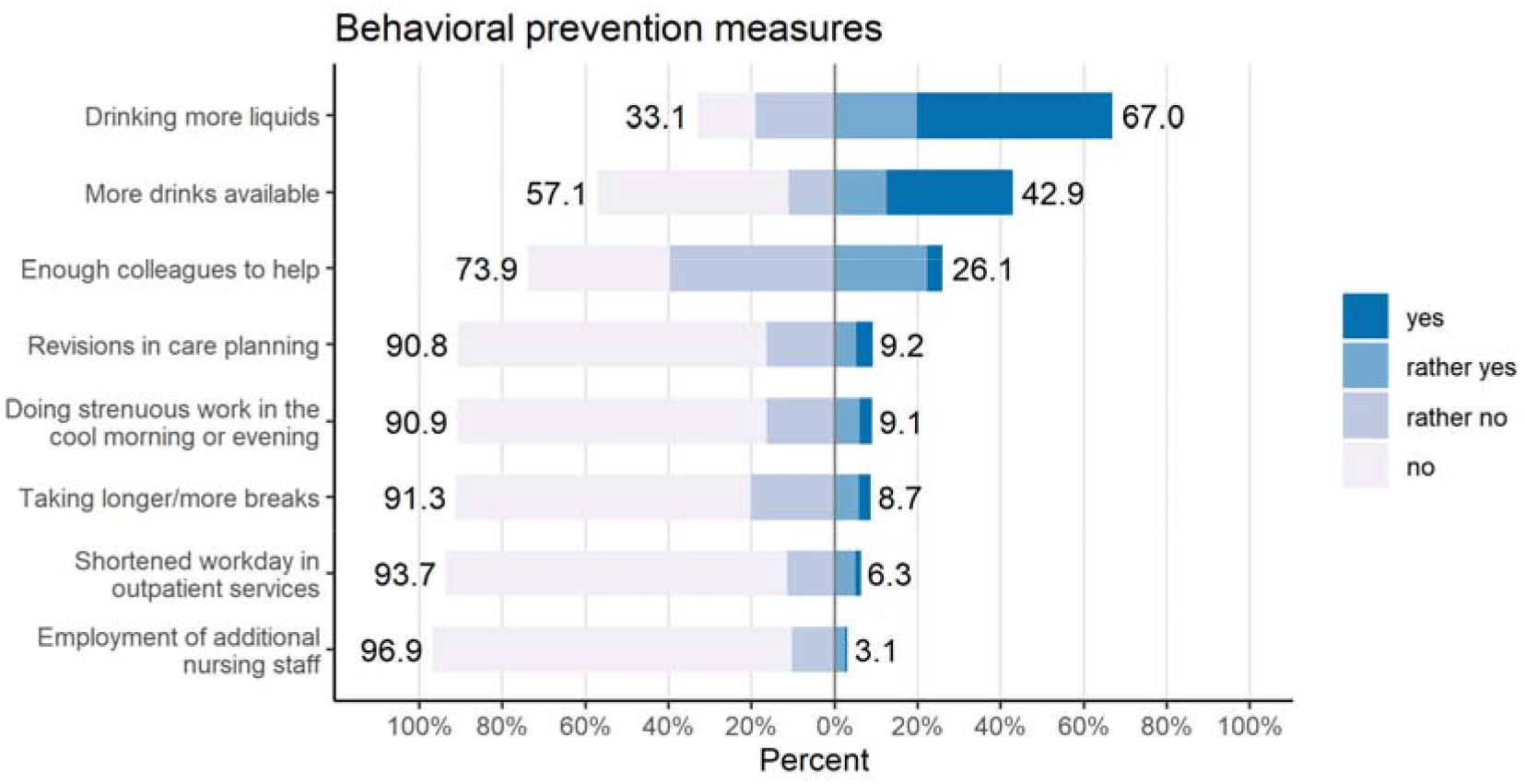
Countermeasures by healthcare workers against excessive heat.

### Heat warnings and heat protection action plans in the facility comparison

55.4% of the nursing staff from the nursing homes said they had received heat warnings and about 22.2% mentioned they knew about heat protection action plans. As a result, 12.5% of the nursing staff felt well protected in the nursing homes. In outpatient care, 24.1% received heat warnings and 6.7% received heat protection action plans (figure 4, table S4,). 22.2% of the nursing staff felt that they were well protected at work. In the clinics, it was 4.8% of the nursing staff who received heat warnings and 0.9% who was aware of heat protection action plans. As a result, 0% of the nursing staff in the hospitals felt well protected. PPE was not always available; while only 7.8% and 9.6% of survey participants reported that surgical caps and protective gloves were missing, respectively, FFP2 and FFP3 masks were absent in 33.7% and 30.0% of cases (figure S2 and S3).

**Figure 4.**
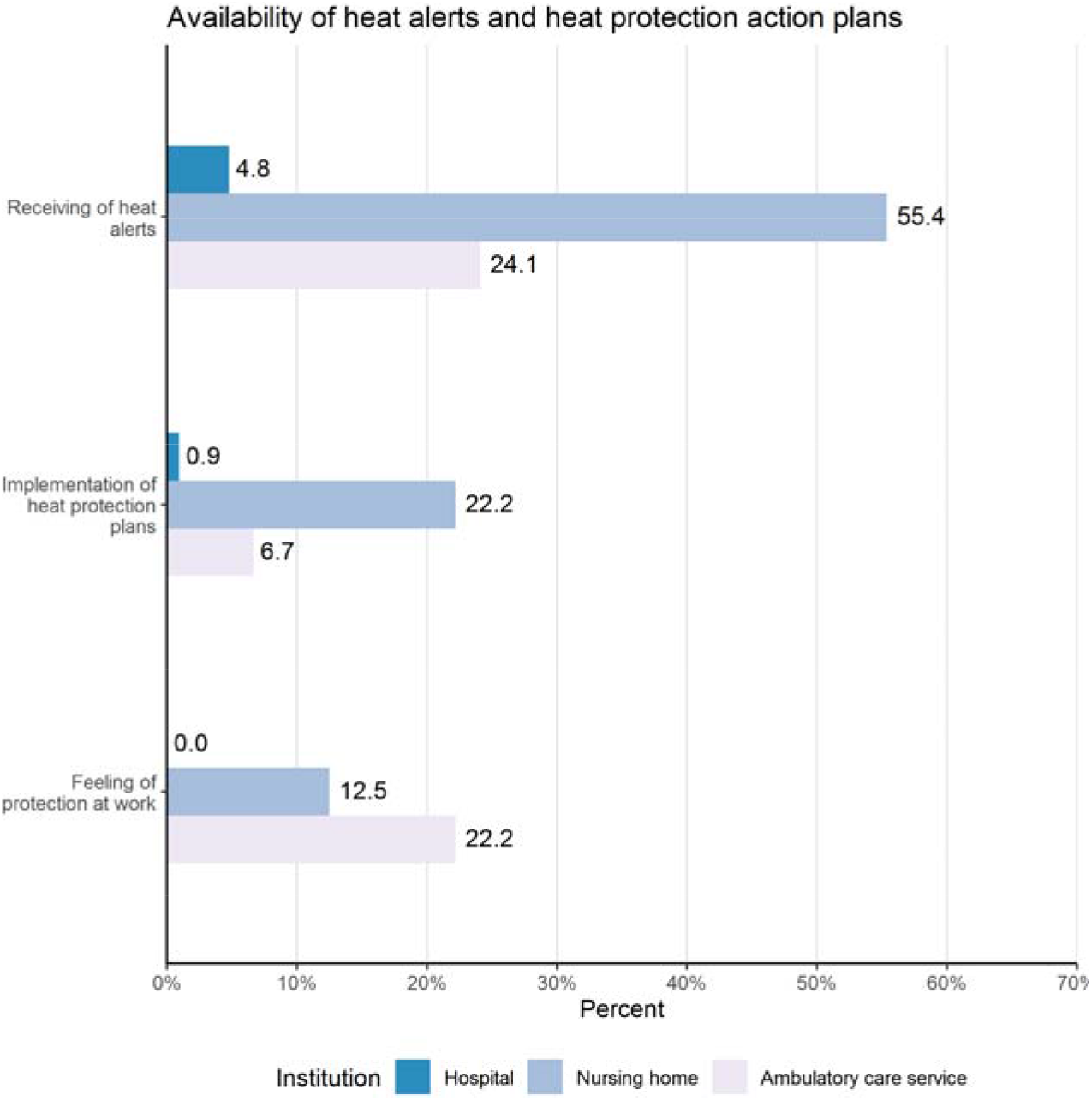
Reception of heat alerts and heat action plans across institutions.

### Condition-oriented prevention when working in PPE on hot days in a comparison of facilities

Figure 5 shows that in nursing homes (55.4%) and in ambulatory care services (57.4%), more drinks were served than in hospitals (33.8%), (p<0.001). In 56.3% of nursing homes, room thermometers were available. This was significantly more than in clinics (29.0%) or ambulatory care (16.7%), (p<0.001). In 59.7% of the nursing homes, refrigerators were available; these numbers were 43.4% in the clinic, and 41.4% in the outpatient care service (p=0.5). The personnel situation was similarly poor in all areas, e.g. 75.1% in the clinic, 66.2% in the home and 81.4% in outpatient care said that there was insufficient staff (p=0.13). Changes in care planning were not made in any of the settings (87.7-92.8%) (p=0.3). See also figure S1 for behaviour-oriented preventive measures.

**Figure 5.**
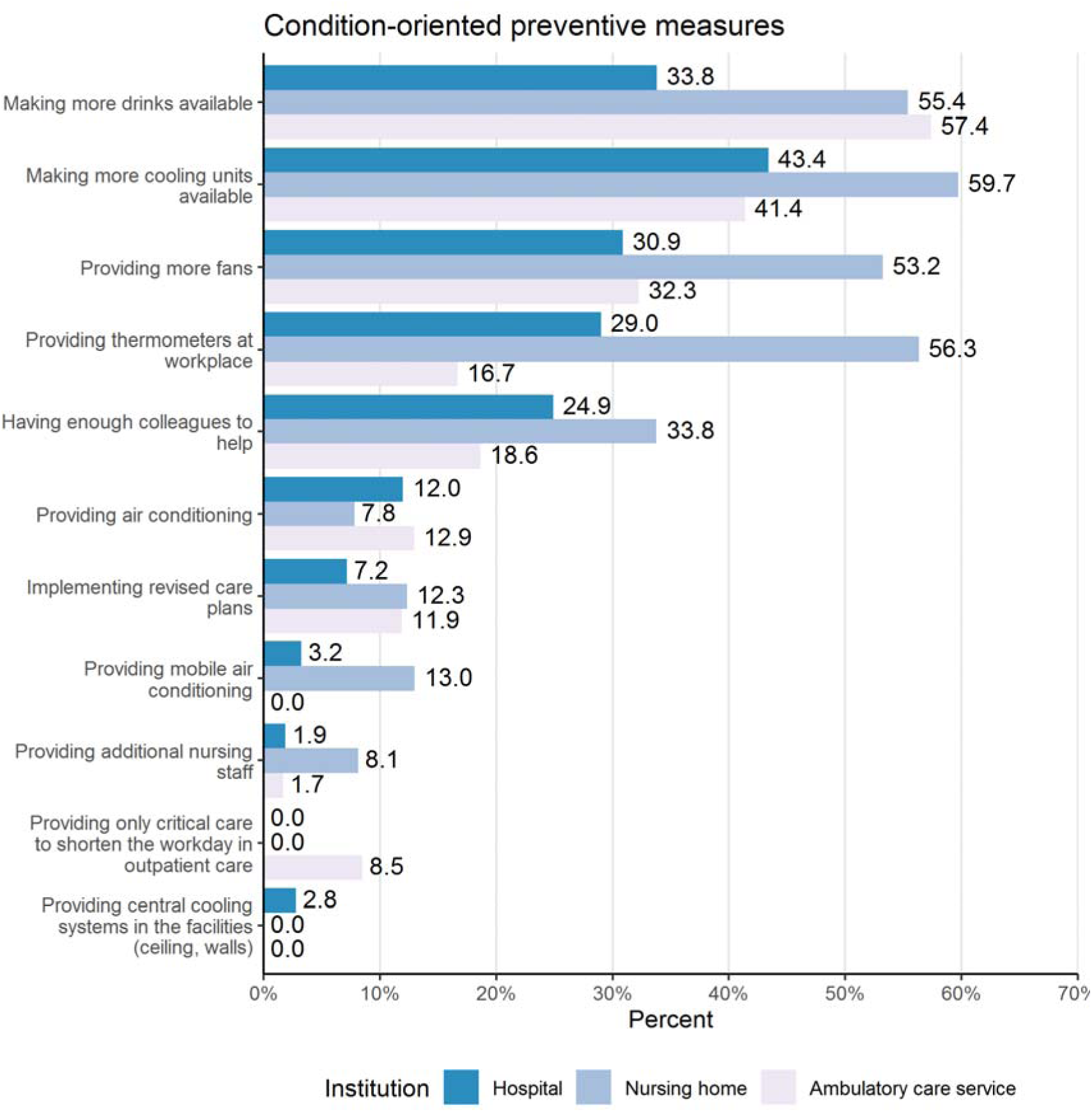
Condition-oriented preventative measures against heat stress.

Permanently installed air conditioning systems were available in hospitals, as reported by 12% of nursing staff; in nursing homes, it was 7.8%, and 12.9% in ambulatory care. These differences were statistically not significant with p=0.5. Of all participants, 13.0% mentioned the use of mobile cooling devices in nursing care facilities. The clinic had 3.2% of these devices, and 0.0% of them were available in the context of outpatient care (p<0.001). In nursing homes, 53.2% used fans, 30.9% in clinics and 32.3% in outpatient care (p=0.002). There were no more fixed air conditioning systems in the hospitals than in other settings (figure 5).

## Discussion

In our survey, the majority of nurses and healthcare workers was female, and a plurality was over 45 years of age. In most cases, these professionals had also more than 10 years of experience, often 20 years. This is in line with the public understanding of the nursing field being dominated by women; however, the lack of younger nurses could also be due to a perceived lack of appreciation and low pay [18].

Two thirds of participants (59%) felt that their work was not appreciated, which may have contributed to feelings of overwhelm and psychological or even physical hardship. With 91% the vast majority of individuals taking part in the survey suffered from various complaints like back pain, headaches, exhaustion, sleep deprivation and so forth. In this context it is important to mention that almost half of all participants (46%) already suffered from pre-existing conditions like hypertension, depression, diabetes and other. These problems could become enhanced when being subjected to mentally and physically taxing work with uncomfortable clothing in hot temperatures.

Working in PPE during hot days increased respondents’ discomfort; almost everyone complained from more sweating and shortness of breath, and half of the participants were afraid to make mistakes and became stressed when changing PPE during shifts. This matches with previously published results [19] and suggests that the anguish and discomfort felt when wearing PPE was both due to wearing the clothing and the elevated temperatures. Recent studies have pointed out that discomfort and stress increased with the duration of wearing PPE. This suggests that the stress felt while wearing PPE is not just a psychological effect but linked to actually wearing the clothing [20]. Irrespective of which possibility applies, the survey results suggest that healthcare organizations should provide opportunities to physicians and nursing staff to alleviate the heat stress they experience, including institutional policies outlining a maximum duration for which PPE could be worn, at least during peak temperatures [20].

Offering opportunities and implementing the right policies appears important also because in our study, the overwhelming majority of nurses and healthcare workers were not able to effectively deploy countermeasures against heat stress on their own. As two thirds of study participants (67%) reported that they drank more water, it may make sense for healthcare centres to offer extra water or ice slurries, as has been recommended in one recent study [9]. Offering cold water temperatures has both a cooling effect and increases the likelihood that nurses take up more fluid during their shifts. Moreover, training staff to frequently drink water will not only help during heat waves but will in general work against another common danger in the workplace - dehydration [9]. One possibility to incorporate this into the nursing shifts would be for the employer or hospital to inform nurses of this option and offering boxes filled with ice for everyone to use. Another way to mitigate heat stress would be to offer longer breaks to nurses, so they have more time to recuperate [21]. This can be surprisingly difficult to implement, as healthcare workers may feel the need to treat patients while they are in the hospital, especially if the hospital is understaffed, as was reported by almost every participant of our survey; this is consistent with previously published research [22]. Introducing more rest breaks may not only help provide heat relief, but also mitigate workplace stress. Therefore, having mandatory break times may help reduce with nursing staff turnover, which would be a distinct advantage especially during pandemics like SARS-CoV-2 [23]. A large number of care facilities participants of our questionnaire worked in did not have a clear heat action plan, and even simple devices like room thermometers and additional refrigerators were missing. In cases where it is difficult to implement air conditioning, mobile cooling devices can help; less than a fifth of hospitals, nursing homes and ambulatory care settings had those available at the time of our survey. Foster et al. recommend a combination of ingestion of water and ice slurries, air conditioning - if financially possible for the corresponding institution - and shading in areas that are strongly impacted by heat [9].

In summary, there is a distinct need for better institutional policies that acknowledges the existence of heat stress and the need of cooling for nursing personnel and healthcare workers. This is especially true as climate change will not only increase average temperatures in the future, but also the number of heat waves that occur within a year [3].

The results of our study suggest that adequate heat mitigation steps should be taken by hospitals, care homes and even in ambulatory care, to protect the nursing staff from heat stress in combination with, or exacerbating, the wearing of PPE. Institutional recommendations are not guaranteed to be universally applied. On the long-term, it may be necessary to add heat protection measures for healthcare workers to occupational health and safety laws. Importantly, such laws need to be stringently enforced, as even Germany - which already has such occupational safety laws in place, whereby air temperatures indoor cannot be higher than 26 °C, and companies have to provide adequate heat protection for their employees - still has nursing staff suffering from heat stress, as shown in this study [24]. Such resolutions need to come from the legislative level; it is important to note that despite the systemic relevance of nursing staff during the Covid-19 pandemic, concrete political steps to protect healthcare workers have, so far, been scarce [25]. The expected increase in future demand for the nursing profession suggests that there are good reasons to improve working conditions for healthcare workers. For example, the growth in professional positions for registered nurses in Germany and the United States is faster than average, with the projected growth in residential care facilities even larger [26]. Thus, improving working conditions for nurses will become more and more important and will likely include heat protection measures for nurses that have to wear PPE [27]. It is not entirely clear whether modest pay raises for nursing staff alone, as recently implemented in Germany, will be a viable long-term solution [28].

As countries in Southeast Asia have been among the first ones being exposed to the Covid-19 pandemic, it may be instructive to look at the well-being of healthcare workers in those nations, and how they have dealt with wearing PPE in hot climates. Studies show that the psychological burden among nurses in Vietnam, Malaysia, Singapore, India and Indonesia has been largely independent of the SARS-CoV-2 case load in those countries, and instead depends on the level of medical training, the presence of physical symptoms and prior medical conditions. Thus, psychological adversity is not just a temporary outcome of the pandemic burden but has deeper roots [29]. These observations also support the notion that any measures taken to improve heat protection among nurses working in PPE is of more general importance.

Another survey among Indian and Singaporean healthcare workers shows similar sentiments to the results in this study. 75% of nurses found that wearing of PPE made them uncomfortable, and 47% even found that their productivity was reduced. Most nurses wore N95 or surgical masks as well as gloves and gowns. Importantly, the ingestion of ice slurries significantly improved Singaporean HCWs heat comfort, suggesting that it may be a viable method of heat protection for nurses working in PPE [5].

### Limitations

Our survey was not designed to address whether temperature and ‘clothing discomfort’ applied independently of one another, or whether the elevated temperature exacerbated existing discomfort from wearing PPE. One way of distinguishing both possibilities would be a randomized control study with four groups of nurses, that would or would not wear PPE and are exposed to normal or hot room temperature. The limitations of this study are imbalances in the sample (distribution of registered nurses, nursing assistants, geriatric nurses) and the possibility of a selection bias in favour of the participation of dissatisfied older employees. Moreover, the survey was done exclusively with nurses in Germany. Selection bias is also possible due to the participation of nursing professionals associated with professional associations and online publishers; we did not assess from which geographic region the participants came. This group of participants may epitomize employees with special characteristics that are therefore not representative of healthcare workers in general.

## Conclusions

Overall heat and wearing of PPE both causes discomfort in nurses and healthcare workers. It is important that the employers have a strong strategy in place to mitigate negative effects from heat, especially since future demand for nurses will go up; such a strategy could include training for nurses to inform them of the necessity of protection from heat and the existing possibilities. Moreover, hospitals, nursing homes and outpatient services should offer several tools to reduce heat and associated stress, such as air conditioning or cooling devices, ice-water baths for consumption, and an updated break schedule that allows nurses to properly regain their mental and physical strength during their respective shift. It would be interesting to know whether the heat stress reported by nursing personnel in this study lead to an increased willingness of changing professions, a desire to move to a different healthcare area, or an intention to decrease working hours. The correlation between job satisfaction and choosing to remain working in a particular job does not always appear to be positive for nursing staff, at least not in Germany [30]. Future studies should assess the impact of heat protection measures and the wearing of PPE during hot ambient temperatures on job satisfaction and turnover.

## Data Availability

data are available on resonable request

## Notes

**Conflict of interest statement for all authors** The authors declare that there is no conflict of interest regarding the publication of this paper.

### Competing Interest Statement

The authors have declared no competing interest.

### Funding Statement

Funded by the Federal Ministry for the Environment, Nature Conservation and Nuclear Safety (BMU) on the basis of a resolution of the German Bundestag (project number: 67DAS213)

