## Supplementary material for "Hot days and Covid-19 – unusual heat stress for nursing professions in Germany": Suppl

### Supplement

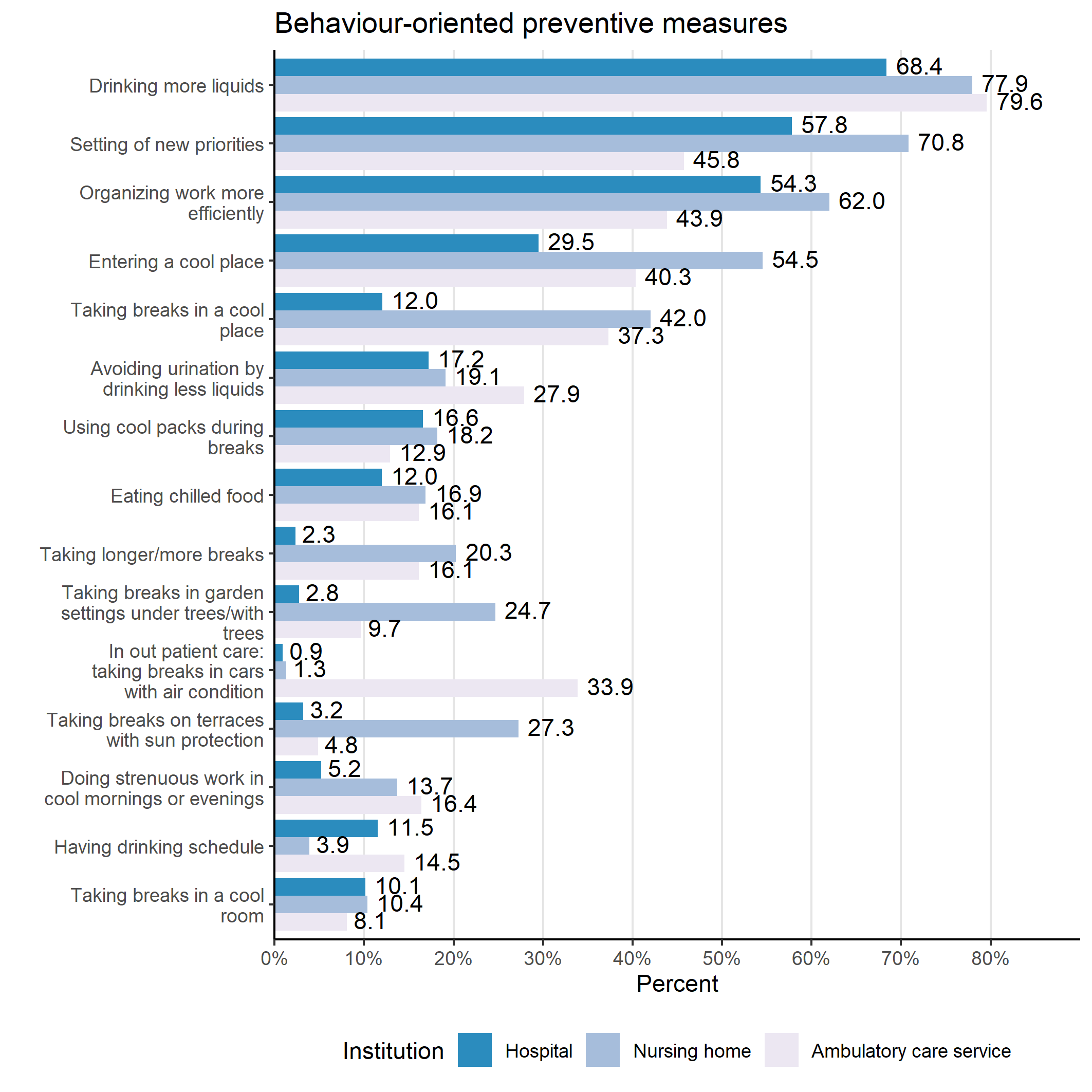

**Figure S1**. Behavioural measures to prevent and mitigate heat stress and their application across different institutions.

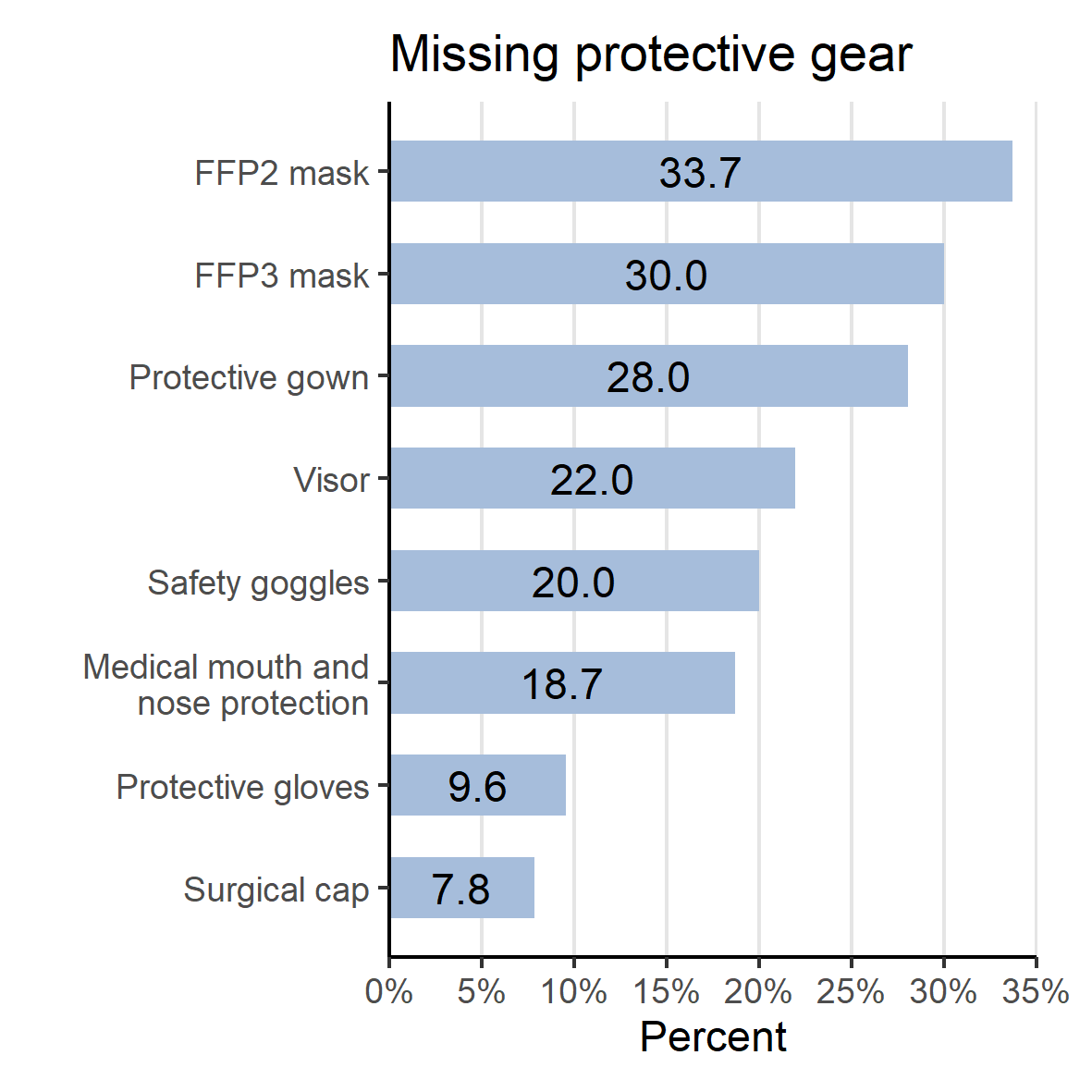

**Figure S2**. Amount in which protective gear is missing from healthcare environments.

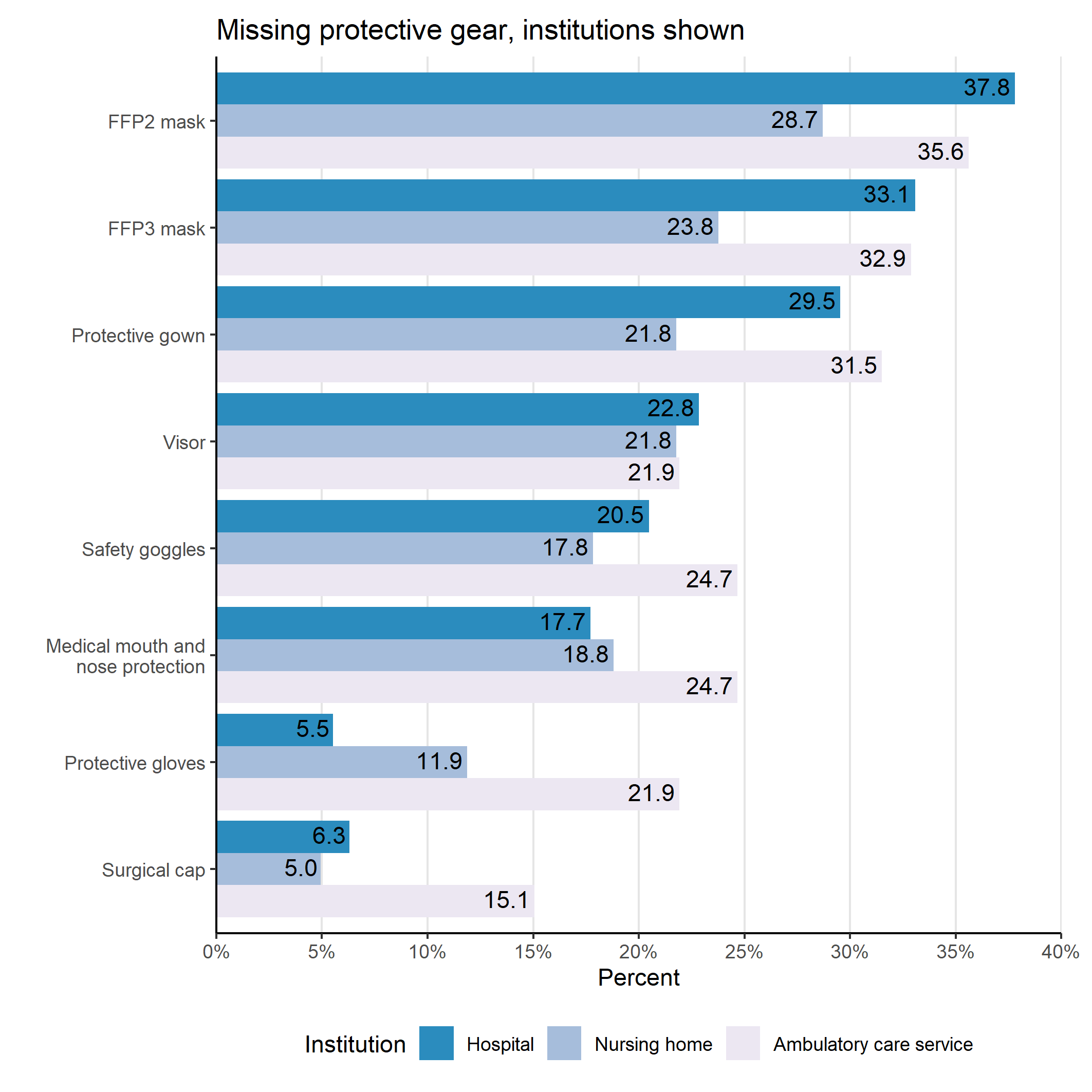

**Figure S3**. Amount in which protective gear is missing from healthcare environments across different institutions.

### Tables

|  | Institution | | |  |
| --- | --- | --- | --- | --- |
| Characteristics | Hospital, N = 254^1^ | Nursing home, N = 101^1^ | Ambulatory care service, N = 73^1^ | Total, N = 428^1^ |
| Gender |  |  |  |  |
| Male | 43 (20.1%) | 14 (20.0%) | 4 (6.5%) | 61 (17.6%) |
| Female | 171 (79.9%) | 56 (80.0%) | 58 (93.5%) | 285 (82.4%) |
| Age |  |  |  |  |
| 16-25 years | 20 (9.3%) | 2 (2.9%) | 0 (0.0%) | 22 (6.3%) |
| 26-35 years | 57 (26.5%) | 10 (14.3%) | 12 (19.4%) | 79 (22.8%) |
| 36-45 years | 44 (20.5%) | 20 (28.6%) | 12 (19.4%) | 76 (21.9%) |
| 45-55 years | 63 (29.3%) | 24 (34.3%) | 19 (30.6%) | 106 (30.5%) |
| 56-65 years | 30 (14.0%) | 14 (20.0%) | 19 (30.6%) | 63 (18.2%) |
| 65+ | 1 (0.5%) | 0 (0.0%) | 0 (0.0%) | 1 (0.3%) |
| Working hours |  |  |  |  |
| Full-time | 125 (58.7%) | 48 (69.6%) | 40 (64.5%) | 213 (61.9%) |
| Part-time | 86 (40.4%) | 20 (29.0%) | 22 (35.5%) | 128 (37.2%) |
| Mini job | 2 (0.9%) | 1 (1.4%) | 0 (0.0%) | 3 (0.9%) |
| Employment relationship |  |  |  |  |
| Directly employed | 207 (96.3%) | 68 (98.6%) | 57 (91.9%) | 332 (96.0%) |
| Leasing | 6 (2.8%) | 1 (1.4%) | 1 (1.6%) | 8 (2.3%) |
| Self-employed | 2 (0.9%) | 0 (0.0%) | 4 (6.5%) | 6 (1.7%) |
| Duration of employment |  |  |  |  |
| < 1 year | 7 (3.3%) | 2 (2.9%) | 2 (3.2%) | 11 (3.2%) |
| 1-2 years | 4 (1.9%) | 3 (4.3%) | 2 (3.2%) | 9 (2.6%) |
| 3-5 years | 20 (9.4%) | 9 (12.9%) | 2 (3.2%) | 31 (9.0%) |
| 6-10 years | 38 (17.9%) | 7 (10.0%) | 10 (16.1%) | 55 (16.0%) |
| 11-20 years | 41 (19.3%) | 20 (28.6%) | 11 (17.7%) | 72 (20.9%) |
| > 20 years | 102 (48.1%) | 29 (41.4%) | 35 (56.5%) | 166 (48.3%) |
| Previous illnesses | 111 (44.0%) | 49 (48.5%) | 36 (50.7%) | 196 (46.2%) |
| Working mostly alone | 61 (24.4%) | 41 (40.6%) | 66 (91.7%) | 168 (39.7%) |
| Enough protective clothing at workplace | 176 (69.8%) | 68 (67.3%) | 49 (67.1%) | 293 (68.8%) |
| Hygiene plan at workplace | 232 (98.3%) | 93 (96.9%) | 65 (97.0%) | 390 (97.7%) |
| Physically demanding work | 236 (93.7%) | 91 (91.0%) | 60 (83.3%) | 387 (91.3%) |
| Feeling valued | 78 (31.1%) | 53 (53.0%) | 41 (57.7%) | 172 (40.8%) |
| Frequently suffering from physical complaints | 206 (83.1%) | 77 (77.8%) | 53 (73.6%) | 336 (80.2%) |
| ^1^Measures: n (%) | | | | |

**Table S1**. Participants’ demographics and information about employment.

| Characteristics | Hospital, N = 254^1^ | Nursing home, N = 101^1^ | Ambulatory care service, N = 73^1^ | p-value^2^ |
| --- | --- | --- | --- | --- |
| Taking longer/more breaks | 5 (2.3%) | 15 (20.3%) | 10 (16.1%) | <0.001 |
| Drinking more liquids | 129 (60.3%) | 60 (81.1%) | 46 (74.2%) | 0.002 |
| Doing strenuous work in the cool morning or evening | 11 (5.2%) | 10 (13.7%) | 10 (16.4%) | 0.007 |
| Setting of new priorities | 115 (57.8%) | 51 (70.8%) | 27 (45.8%) | 0.014 |
| Organizing work more efficiently | 113 (54.3%) | 44 (62.0%) | 25 (43.9%) | 0.12 |
| Avoiding urination by drinking less liquids | 36 (17.2%) | 13 (19.1%) | 17 (27.9%) | 0.2 |
| Drinking more | 132 (68.4%) | 53 (77.9%) | 39 (79.6%) | 0.15 |
| Drinking schedule | 25 (11.5%) | 3 (3.9%) | 9 (14.5%) | 0.086 |
| Eating chilled food | 26 (12.0%) | 13 (16.9%) | 10 (16.1%) | 0.5 |
| Using cool packs during breaks/ cold compresses | 36 (16.6%) | 14 (18.2%) | 8 (12.9%) | 0.7 |
| Entering cooler place | 64 (29.5%) | 42 (54.5%) | 25 (40.3%) | <0.001 |
| Taking breaks in cool place | 25 (12.0%) | 29 (42.0%) | 22 (37.3%) | <0.001 |
| Breaks in cool room | 22 (10.1%) | 8 (10.4%) | 5 (8.1%) | 0.9 |
| Breaks in garden under trees | 6 (2.8%) | 19 (24.7%) | 6 (9.7%) | <0.001 |
| Breaks on terrace with sun protection | 7 (3.2%) | 21 (27.3%) | 3 (4.8%) | <0.001 |
| In outpatient care: taking breaks in car with air conditioning | 2 (0.9%) | 1 (1.3%) | 21 (33.9%) | <0.001 |
| ^1^Measurements: n (%) | | | | |
| ^2^Statistical tests: Chi-square-test; Fisher’s exact test | | | | |

**Table S2**. Heat avoiding behaviours among participants.

| Characteristics | Hospital, N = 254^1^ | Nursing home, N = 101^1^ | Ambulatory care service, N = 73^1^ | p-value^2^ |
| --- | --- | --- | --- | --- |
| Making more drinks available | 71 (33.8%) | 41 (55.4%) | 35 (57.4%) | <0.001 |
| Enough colleagues to help | 53 (24.9%) | 25 (33.8%) | 11 (18.6%) | 0.13 |
| Providing additional nursing staff | 4 (1.9%) | 6 (8.1%) | 1 (1.7%) | 0.040 |
| Implementing revised care plans | 15 (7.2%) | 9 (12.3%) | 7 (11.9%) | 0.3 |
| Providing only critical care to shorten workday in outpatient services | 0 (0.0%) | 0 (0.0%) | 5 (8.5%) | 0.7 |
| Providing thermometers at workplace | 58 (27.2%) | 40 (54.8%) | 9 (15.3%) | <0.001 |
| Making more cooling units available | 89 (42.2%) | 43 (58.9%) | 24 (41.4%) | 0.037 |
| Providing air conditioning | 26 (12.0%) | 6 (7.8%) | 8 (12.9%) | 0.5 |
| Providing mobile air conditioning | 7 (3.2%) | 10 (13.0%) | 0 (0.0%) | <0.001 |
| Providing central cooling systems in the facilities (ceiling, walls) | 6 (2.8%) | 0 (0.0%) | 0 (0.0%) | 0.3 |
| Providing fans | 67 (30.9%) | 41 (53.2%) | 20 (32.3%) | 0.002 |
| ^1^Measurements: n (%) | | | | |
| ^2^ Statistical tests: Chi-square-test; Fisher’s exact test | | | | |

**Table S3**. Heat mitigation measures.

|  | Heat warnings | | | | HPAP known | | | | HPAP protects | | | |
| --- | --- | --- | --- | --- | --- | --- | --- | --- | --- | --- | --- | --- |
| Characteristics | Yes | No | Do not know | Total | Yes | No | Do not know | Total | Yes | No | Do not know | Total |
| **Institution** |  |  |  |  |  |  |  |  |  |  |  |  |
| Hospital | 10 (4.8%) | 191 (91.0%) | 9 (4.3%) | 210 (100.0%) | 2 (0.9%) | 202 (95.3%) | 8 (3.8%) | 212 (100.0%) | 0 (0.0%) | 32 (68.1%) | 15 (31.9%) | 47 (100.0%) |
| Nursing home | 41 (55.4%) | 29 (39.2%) | 4 (5.4%) | 74 (100.0%) | 16 (22.2%) | 49 (68.1%) | 7 (9.7%) | 72 (100.0%) | 4 (12.5%) | 18 (56.2%) | 10 (31.2%) | 32 (100.0%) |
| Ambulatory care service | 14 (24.1%) | 39 (67.2%) | 5 (8.6%) | 58 (100.0%) | 4 (6.7%) | 51 (85.0%) | 5 (8.3%) | 60 (100.0%) | 4 (22.2%) | 10 (55.6%) | 4 (22.2%) | 18 (100.0%) |
| No answer | 0 (NA%) | 0 (NA%) | 0 (NA%) | 0 (NA%) | 0 (NA%) | 0 (NA%) | 0 (NA%) | 0 (NA%) | 0 (NA%) | 0 (NA%) | 0 (NA%) | 0 (NA%) |
| **Total** | 65 (19.0%) | 259 (75.7%) | 18 (5.3%) | 342 (100.0%) | 22 (6.4%) | 302 (87.8%) | 20 (5.8%) | 344 (100.0%) | 8 (8.2%) | 60 (61.9%) | 29 (29.9%) | 97 (100.0%) |

**Table S4**. Availability of heat warnings and heat-prevention-action-plans.
